# Navigating the Crisis: A Cross-sectional Survey Analysis of Resident Doctors’ Experiences of Specialty Training and Employment in Today’s NHS

**DOI:** 10.64898/2025.12.07.25341779

**Authors:** Barbara Dorgan, Rachel Elliott, Stephen J. Dorgan

## Abstract

**Objective:** To investigate training conditions, career opportunities, workforce perceptions, and the emotional strain encountered by resident doctors within the NHS’s current training and recruitment pathways.

**Design:** A cross-sectional, anonymous survey of UK resident doctors conducted via an online questionnaire. Quantitative and qualitative data on employment status, training opportunities, recruitment processes, and emotional wellbeing, from both the anonymous survey and NHS datasets, were collected and analysed via descriptive statistics and thematic analysis.

**Setting:** The NHS faces a deepening crisis. The demand for doctors continues to rise, yet qualified resident doctors encounter increasing barriers to entering specialty training or gaining secure employment.

**Participants:** A total of 288 UK resident doctors received the survey in June and July 2025. Participation was voluntary, and no incentives were offered. 102 surveys were completed, giving an overall response rate of 35%.

**Results:** Respondents reported growing competition for both specialty training and local employment posts, which is supported by NHS training and employment data. Only 39% of respondents secured Specialty or Higher Training post, while just 8% were successful applying for a trust-grade position despite substantial time and effort invested. Two-thirds (66%) reported missing out on advertised roles due to recruitment system failures. Respondents reported overwhelmingly negative emotions relating to fear, sadness and anger which directly impact staff morale and wellbeing. This manifests in significant stress, anxiety, and dissatisfaction with their career prospects alongside feelings of hopelessness, disillusionment, and demotivation. Over one-fifth (21%) have left the NHS or medicine altogether over the last two years.

**Conclusions:** Resident doctors currently face unprecedented challenges in applying to, and progressing in, NHS training and recruiting systems. These difficulties are having a profound effect upon morale, wellbeing, and career viability, which is contributing to increased attrition from both the NHS and the medical profession. Urgent reforms to both training pathways and recruitment processes are needed to safeguard both workforce retention and high-quality patient care.

**Article Summary:** *Strengths and limitations of this study:*

- This study provides timely evidence on resident doctors’ experiences of current NHS training and employment pathways during a period of unprecedented competition.
- A mixed-methods approach, combining survey data with qualitative analysis and publicly available NHS workforce datasets, strengthens the depth and contextual relevance of the findings.
- Selection bias is possible, as doctors with negative experiences may have been more likely to participate.
- The focus on early-career doctors restricts the applicability of the findings to higher specialty training contexts.

## Introduction

In recent years, the landscape for specialty medical training and employment of resident doctors in the NHS has shifted dramatically. Despite growing evidence of the urgent need to expand the medical workforce, many resident doctors are unable to progress in their careers and access training opportunities or secure local employment to meet service needs.

The NHS remains under severe strain. Over 7.3 million patients remain on NHS waiting lists [1], and public satisfaction with the NHS is at its lowest level in over 40 years [2]. Persistent rota gaps leave tens of thousands of shifts unfilled [3], and workforce burn-out is increasingly common [4]. While there is both a growing supply and a clear demand for more doctors, the current employment market structures appear unable of matching the supply of doctors with the demand.

A key driver of the mismatch is the shortage of training and non-training posts for resident doctors. While applications to Specialty Training have increased almost 300% since 2019, the number of training positions have increased by only 5% over the same period [5]. This imbalance has led to widespread frustration, with some groups, such as Foundation Year 1 (FY1) doctors, voting for industrial action in response to the lack of available positions to progress their careers [6].

Relatively recent policy changes have further compounded the issue. The removal of the Resident Labour Market Test (RLMT) and the expansion of UK medical school output [7] sought to address a medical workforce shortage across the NHS. However, without a dramatic and rapid increase in specialty training capacity these two policy changes risk exacerbating the situation, worsening both competition and attrition amongst resident doctors wishing to work in the NHS and progress in their careers.

This study presents findings from a cross-sectional survey conducted in mid-2025. It examines the experiences, challenges, and perceptions of resident doctors navigating specialty training and longer-term employment pathways within the NHS.

## Methods

### Survey Design and Distribution

A 17-item online questionnaire (*Online Supplemental Appendix Table A3*) was developed to collect both quantitative and qualitative data. The survey included closed questions and free-text comment sections to capture the experiences and perceptions of resident doctors at different stages of their training and employment journey. Participants included UK and international medical graduates, both within and outside formal training programmes, working in the UK or overseas.

The survey link was distributed via WhatsApp groups used by Foundation Programme doctors and various clinical teams. A total of 288 recipients received the survey between 10 June and 20 July 2025. Participation was voluntary, and no incentives were offered.

### Data collection and analysis

Quantitative survey responses were analysed descriptively. Free-text comments were examined using an inductive thematic analysis approach to identify recurring patterns, sentiments, and emerging themes. Two researchers independently reviewed responses to enhance consistency, and any discrepancies were resolved through consensus and discussion.

To put the survey data in context, we performed a secondary analysis of publicly available NHS workforce and training data for the period 2019-2025 [8, 9].

### Ethics

All data were collected anonymously. No personally identifiable information was recorded. Respondents were fully informed in advance about the purpose of the study and provided implied consent by voluntarily completing the survey. Ethical approval was not required under NHS Health Research Authority guidance, as the study did not involve patients or access to identifiable health records.

### Guidelines

STROBE reporting guidelines [10] were used when drafting and revising this manuscript and STROBE reporting checklist [11] completed prior to submission (*Online Supplemental Appendix Table A4)*.

## Results

### Respondent Demographics

A total of 102 surveys were completed, giving an overall response rate of 35%. Table 1 summarises survey participants’ demographics and roles at the time of the survey (June/July 2025) and future roles from September 2025, with additional detail provided in *Online Supplemental Appendix Table A1*. Most respondents (80%) were resident doctors who graduated medical school between 2021 and 2023 and completed their Foundation Training Programme between 2023 and 2025. The majority (89%) were graduates of UK medical schools. Overall, 39% expected to hold a formal Specialty Training post in Autumn 2025, with 33% in Postgraduate Specialty Training (an increase from 22% the previous year) and 6% of respondents in Higher Specialty Training, up from 4% in the previous year.

**Table 1:** Respondent characteristics – Country of qualification and role.

|  | June-July 2025 |  | After Sept. 2025 |  |
| --- | --- | --- | --- | --- |
|  | n | % | n | % |
| UK medical school graduates: |  |  |  |  |
| • UK nationals | 83 | 81% | 83 | 81% |
| • Non-UK nationals | 8 | 8% | 8 | 8% |
| Non-UK medical school graduates: |  |  |  |  |
| • UK national | 2 | 2% | 2 | 2% |
| • Non-UK national | 7 | 7% | 7 | 7% |
| No information given | 2 | 2% | 2 | 2% |
| Role: |  |  |  |  |
| • Foundation Year 1 | 3 | 3% | - | - |
| • Foundation Year 2 | 18 | 18% | 3 | 3% |
| • Postgraduate speciality training | 22 | 22% | 34 | 33% |
| • Higher speciality training | 4 | 4% | 6 | 6% |
| • Locally employed doctor | 29 | 28% | 21 | 21% |
| • Locum doctor | 14 | 14% | 17 | 17% |
| • Doctor working abroad | 11 | 11% | 13 | 13% |
| • Non-clinical role | 1 | 1% | 2 | 2% |
| • Full-time student | - | - | 2 | 2% |
| • Other | - | - | 4 | 4% |

**Notes:** N= 102

More than half of respondents (58%) remained outside formal training pathways. 21% reported working as Locally Employed Doctors (down from 28% the previous year), while 17% were working as locum doctors, (up from 14%). Over one in five respondents (21%) reported leaving the NHS altogether, an increase from 12% the previous year. Of the 21% leaving the NHS, 13% were leaving the UK, while 8% indicated they would no longer work as doctors after September 2025.

### Specialty Training Opportunities

Table 2 shows the overall share of respondents applying for Specialty Training increased from 40% in 2024 to 51% in 2025. However, success rates in securing a training role or an interview declined. In 2025, 63% of applicants failed to secure a training position compared with 57% in 2024, and less than half (48%) were invited to interview in 2025, down from 63% the previous year.

**Table 2:** Speciality training applications, success, and failure rates – by year.

| Speciality training | Recruitment Cycle Year |  |  |  |
| --- | --- | --- | --- | --- |
|  | 2024 |  | 2025 |  |
|  | n | % | n | % |
| Total individuals applying for a speciality training role* | 41 | 40% | 52 | 51% |
| Total speciality training applications made in-year | 60 | - | 79 | - |
| Applications made that resulted in: |  |  |  |  |
| • An interview** | 38 | 63% | 38 | 48% |
| • A training post offer** | 26 | 43% | 29 | 37% |
| • No offer of a training post** | 34 | 57% | 50 | 63% |

**Notes:** \*N= 102 in both 2024 & 2025; \*\*N= 60 in 2024 and 79 in 2025

Figure 1 illustrates the widening gap between the number of Speciality Training applications and the number of available posts. Between 2019 and 2025, the number of applications increased by 299% (from 23,040 to 91,999), while the number of training posts rose by only 5% from 12,175 to 12,833 [8].

**Figure 1:**
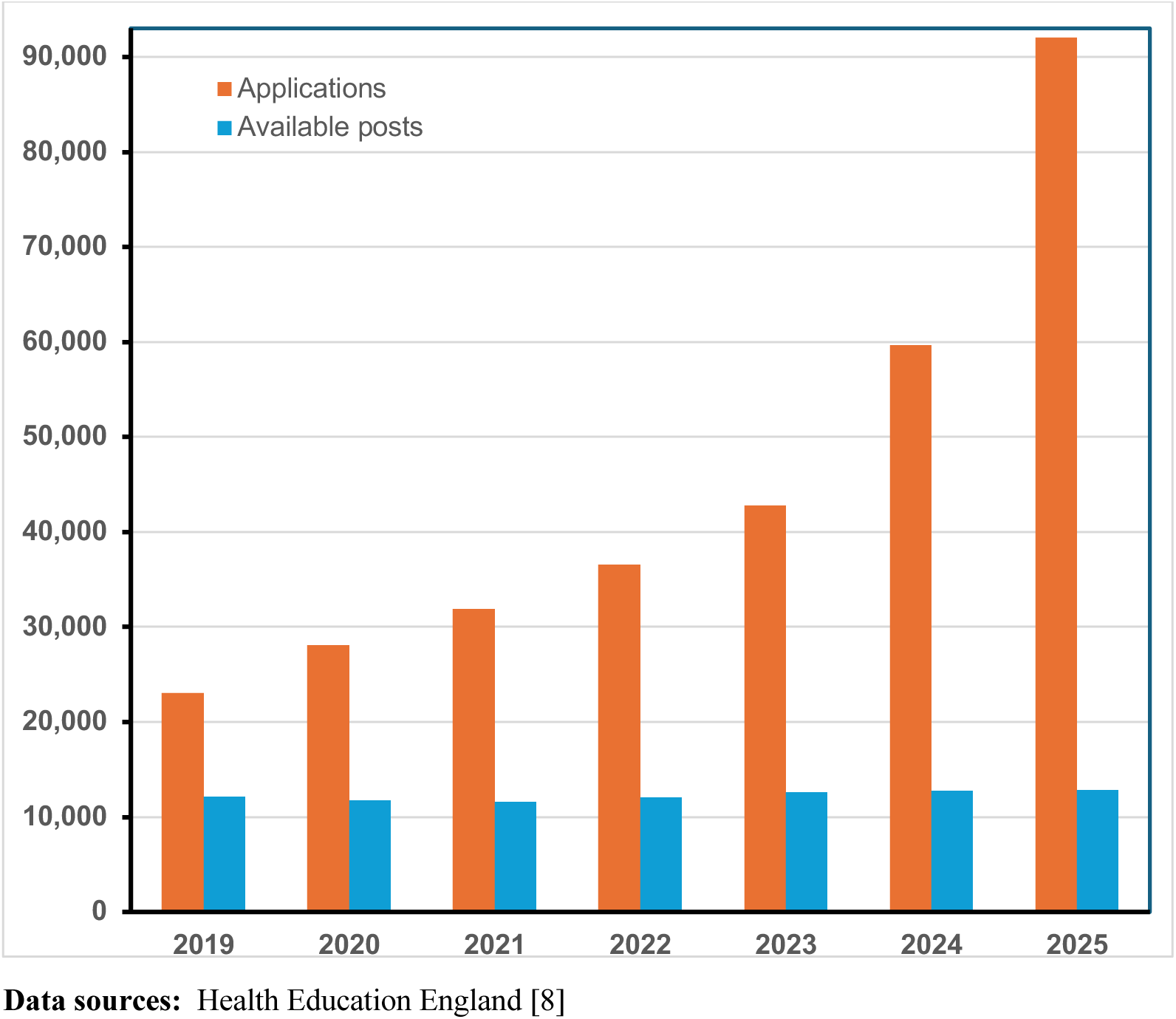
NHS England: Speciality training applications and available posts – 2019-2025.

Figure 2 shows that the recent rise in Specialty Training applications is largely driven by non-UK graduates. Applications from non-UK graduates increased by 208%, from 18,335 in 2021 to 56,453 in 2024, while UK-graduate applications have seen a much more moderate increase of 32%, from 18,195 to 24,068 over the same period [9].

**Figure 2:**
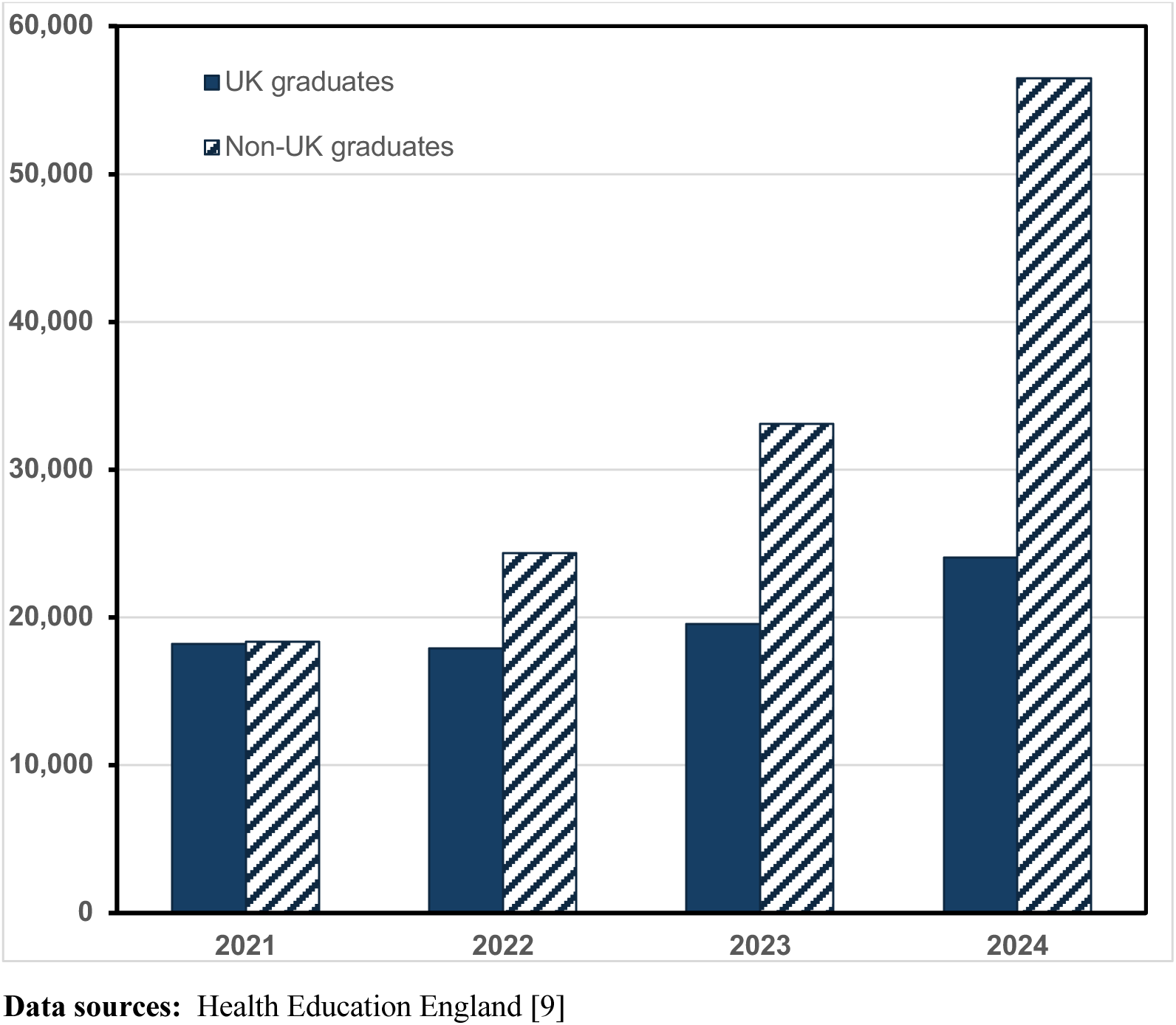
Specialty training applications by country of qualification – 2021-2024.

### Non-Training Employment Opportunities

The secondary impact of limited specialty training post availability upon the wider job market is significant. Over the past two years, 61% of respondents applied for local trust-based roles. Only 15% of these applications led to an interview, and just 8% resulted in a job offer. Overall, 92% of respondents who applied for a local post were unsuccessful in securing any position (*Online Supplemental Appendix Figure A1*).

Competition for these local trust-based roles was intense. Over half of those applying for these trust-based roles (51%) submitted 10 or more applications in 2025, with 3% of respondents submitting over 100 applications. In addition, almost two-thirds (65%) of those applying for trust-based roles reported missing out on an advertised post because the online application window closed prematurely, i.e., ahead of the advertised deadline, preventing them from applying (*Online Supplemental Appendix Figure A2*).

### Satisfaction levels with Training and Job Opportunities

In total, 78% of resident doctors were dissatisfied with current Specialty Training opportunities, even though 39% of the cohort succeeded in securing a Specialty Training post (*Online Supplemental Appendix Figure A3*). This suggests widespread frustration with the effort required and complexity of recruitment processes.

There was similar dissatisfaction with employment outside formal training, with 75% of respondents unhappy with available NHS job opportunities (*Online Supplemental Appendix Figure A4*). Together, these data highlight significant challenges and disillusionment among resident doctors navigating both training and non-training pathways.

### Qualitative Sentiment Analysis

To complement the quantitative data participants were asked: *“Please describe in a few words how the current Postgraduate Medical Specialty Training and wider Resident Doctor job situation in the NHS makes you feel?”* 88 respondents (86% of the total) provided free-text comments, which were analysed thematically. Key themes are summarised in Table 3. Selected illustrative quotes from respondents are also provided in *Online Supplemental Appendix Table A2*.

**Table 3:** Overview of Thematic Insights.

| Key Theme | What doctors report |
| --- | --- |
| Emotional strain and mental health | Feeling broken, emotionally exhausted and on the edge of giving up medicine; suffering deteriorating mental health. |
| Perception of being undervalued and not worthy of investment in future | Feeling undervalued and replaceable, seeing little investment in their future, and viewing medicine as a questionable career choice. |
| Job insecurity and extreme competition | Impossible to secure training posts, unrealistic competition ratios and lack of career progression. |
| Broken by process and ‘survivors’ guilt | Even if successful, doctors feel demotivated and broken by the toll it took, the need to settle for less desirable opportunities and the guilt in seeing other colleagues’ struggles. |
| Systemic issues and mismanagement | A lack of workforce planning, a mismatch of roles and jobs meeting the demand from doctors, prioritisation of service provision over education. |
| Overwhelmed Recruitment systems | Overburdened, failing recruitment systems, not fit for purpose and seen as unfair, untransparent and not selecting for the best candidates. |
| Missed opportunity and inequity | Displacement of local graduates, recruitment not giving sufficient weight to relevant clinical experience, expensive upskilling requirements. |
| Exodus from the NHS and medicine | Intent to leave the NHS or the UK, migrating to countries with better conditions; conflicted about choices and feeling forced into them. |

The qualitative analysis identified a high prevalence of negative emotions with clear implications for morale and wellbeing. Three dominant emotional categories emerged: 1) ‘fear’, reflected in reports of anxiety, worry, insecurity and uncertainty; 2) ‘sadness’, encompassing feelings of hopelessness, despair, being undervalued, and; 3 ) ‘anger’ often intertwined with frustration. Together, these emotions indicate significant psychological distress amongst respondents and appear to contribute to burnout and demoralisation within the workplace, highlighting the significant emotional burden experienced by resident doctors navigating current NHS training and employment pathways.

## Discussion

Our findings demonstrate a clear employment crisis for resident doctors in the NHS, which has been developing over the past six years and is now at a critical point. Three interrelated factors underpin the current situation: (1) the number of Specialty Training posts has remained largely static; (2) applications for training posts have risen sharply; and (3) recruitment processes are overwhelmed and inadequate.

Over half of survey respondents are currently outside formal training roles due to increased competition for those roles. In the 2025 recruitment cycle, 63% of applicants failed to secure any training post, compared with 53% in 2024, and fewer than half (48%) were invited to interview, down from 63% previously. These findings indicate that the selection process is under significant strain, a trend likely to continue unless substantial reforms are implemented.

The lack of training posts is not a new phenomenon. Between 2013 and 2020, there were approximately 2.5 applicants per specialty training post annually [8]. The removal of the Resident Labour Market Test in 2020 contributed to a sharp rise in competition, increasing applicants per post from 1.9 in 2019 to 7.2 in 2025, largely driven by a tripling of applications from non-UK graduates. Non-UK graduates now account for 70% of all applications [9], overwhelming a recruitment system that is unable to process the now 92,000 applications for fewer than 13,000 available posts [5, 8].

Poorly conceived solutions to manage the explosion in training post applications has made the challenge even worse. For example, in Internal Medicine, available training posts have remained static at approximately 1,600 annually since 2015, while interview capacity is limited to around 4,000 [10]. To manage the huge number of applications, interview candidates are selected based upon self-scoring exercises with minimal verification. In fact, applicants are instructed to not provide supporting evidence, unless specifically requested, which they are assured is *“highly unlikely”* [11]. Other Specialty Trainings such as GP Training or Psychiatry have eliminated interviews entirely. These measures, while intended to manage workloads, risk diluting selection rigour and potentially compromising patient care.

The lack of training posts and the growing number of doctors unable to secure Speciality Training roles is increasing competition for non-training roles. More than half of respondents applying for local posts submitted over 10 applications, and some submitted over 100, yet success rates remain low. Fewer than 1 in 12 applicants secured a local role, with inadequate recruitment processes being reported as one of the key drivers of this challenging situation.

Automated systems now prioritise application speed over experience. Two-thirds of all respondents (65%) applying for such local jobs reported missing out on roles because the application window closed earlier than advertised and before they could apply. Increasingly, automated ‘bots’ are immediately applying for roles online as soon as they are advertised despite the applicants being wholly unqualified. In one example, applications for a role closed automatically after just 1.5 hours rather than the advertised 3 weeks, despite 90% of the applications received being substandard. These automated systems effectively exclude highly qualified candidates and undermine any argument that a more competitive labour market will improve candidate quality [12].

These system inefficiencies drive a substantial loss of public investment. The £230,000 taxpayers spend to educate and train each doctor [13] is lost when skilled resident doctors leave the NHS due to poor workforce planning and failing training and employment pathways.

Our survey highlights the broader impact on resident doctors’ morale and wellbeing. Over three quarters of respondents reported dissatisfaction with both Specialty Training and wider job opportunities. Free-text responses reveal pervasive negative emotions of anxiety, stress, hopelessness, and frustration contributing to burnout and low morale. Doctors report struggling with managing both time-consuming application processes and excessive demands by recruiters to demonstrate non-clinical achievements, undermining their ability to recuperate and provide the best patient care.

While policy makers have acknowledged the crisis, and the Chief Medical Officer for England has recommended an overhaul of medical training [14], implementation is unlikely to occur quickly enough to support the current cohort of resident doctors. Without urgent reform, workforce attrition, low morale, and compromised patient care are likely to continue.

## Conclusion

This study highlights a profound and escalating workforce crisis among NHS resident doctors in 2025. Competition for a limited number of Specialty Training posts is intense and increasing, and intensifying competition in the local job market. Doctors who fail to secure a training post often reapply the following year, further increasing training post application volumes. The sharp rise in applications from international medical graduates has intensified competition, resulting in historically high applicant-to-post ratios.

Workforce planning has not kept pace with demand, and recruitment systems are struggling to manage the volume of applications. Current processes are unable to consistently identify the most capable candidates, while measures introduced to cope with high application numbers have inadvertently reduced transparency and fairness. The resulting burden on resident doctors, including excessive extra-curricular expectations, repeated unsuccessful applications, and early elimination from recruitment rounds, has had a detrimental effect on morale, confidence, and mental wellbeing.

These systemic failures are driving widespread dissatisfaction and accelerating the loss of trained talent from the NHS and the wider medical profession. Resident doctors report high levels of anxiety, stress, and burnout, placing both their wellbeing and patient care at risk.

The consequences extend beyond individual doctors: public investment in medical training is being compromised as qualified doctors leave the NHS due to structural inefficiencies and ineffective workforce policies. Greater awareness is needed among policymakers and the public that while patients experience delays in care, a pool of skilled, underutilised resident doctors exists, constrained by system inadequacies rather than capacity or capability. Urgent reforms are required to expand training opportunities, improve recruitment systems, and safeguard both workforce retention and patient care.

*“Retaining talent and fixing the system is not only fair – it’s essential for the future of the NHS.”*

## Data Availability

The data generated during the research and analysis are not available publicly but are available from the corresponding author on a reasonable request.

## Acknowledgments

Thank you to the 102 resident doctors who contributed their time and experience to this survey. A special thank you goes to Dr Catherine Hogel who kindly provided editorial input to an earlier version.

## Author Contributions

BD conceived and designed the study, collected the data, led the primary and secondary data analysis and interpretation, and drafted the first version of the manuscript. RE contributed to the data analysis and context and revised further drafts of the manuscript. SD contributed to the data analysis and critically reviewed further drafts of the manuscript. All authors reviewed and approved the final manuscript.

## Patient and public involvement

Patients and/or the public were not involved in the design, conduct, reporting, or dissemination plans of this research.

## Online Supplemental Appendix

**Figure A1:**
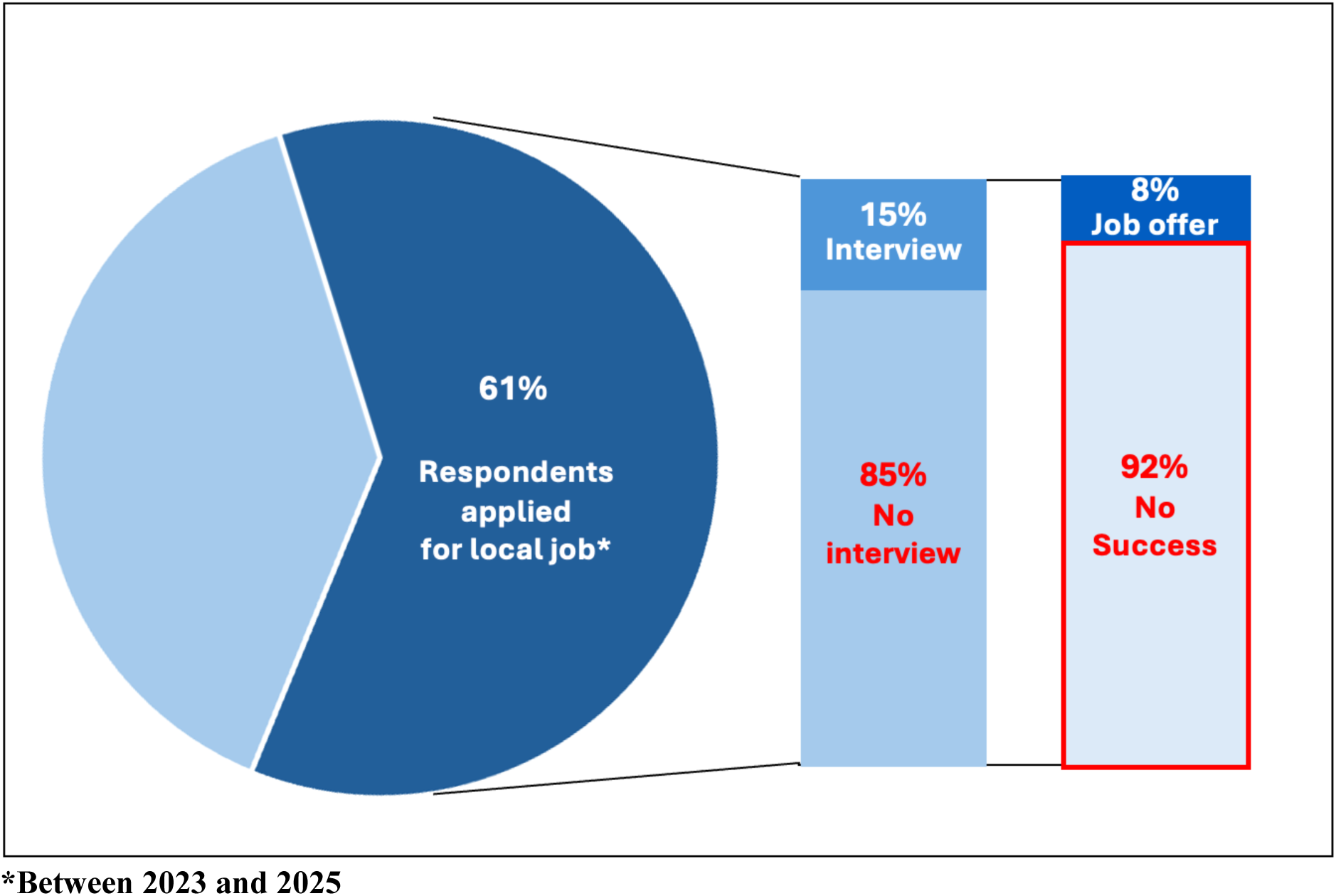
Job Application Success/Failure Rates, 2025.

**Figure A2:**
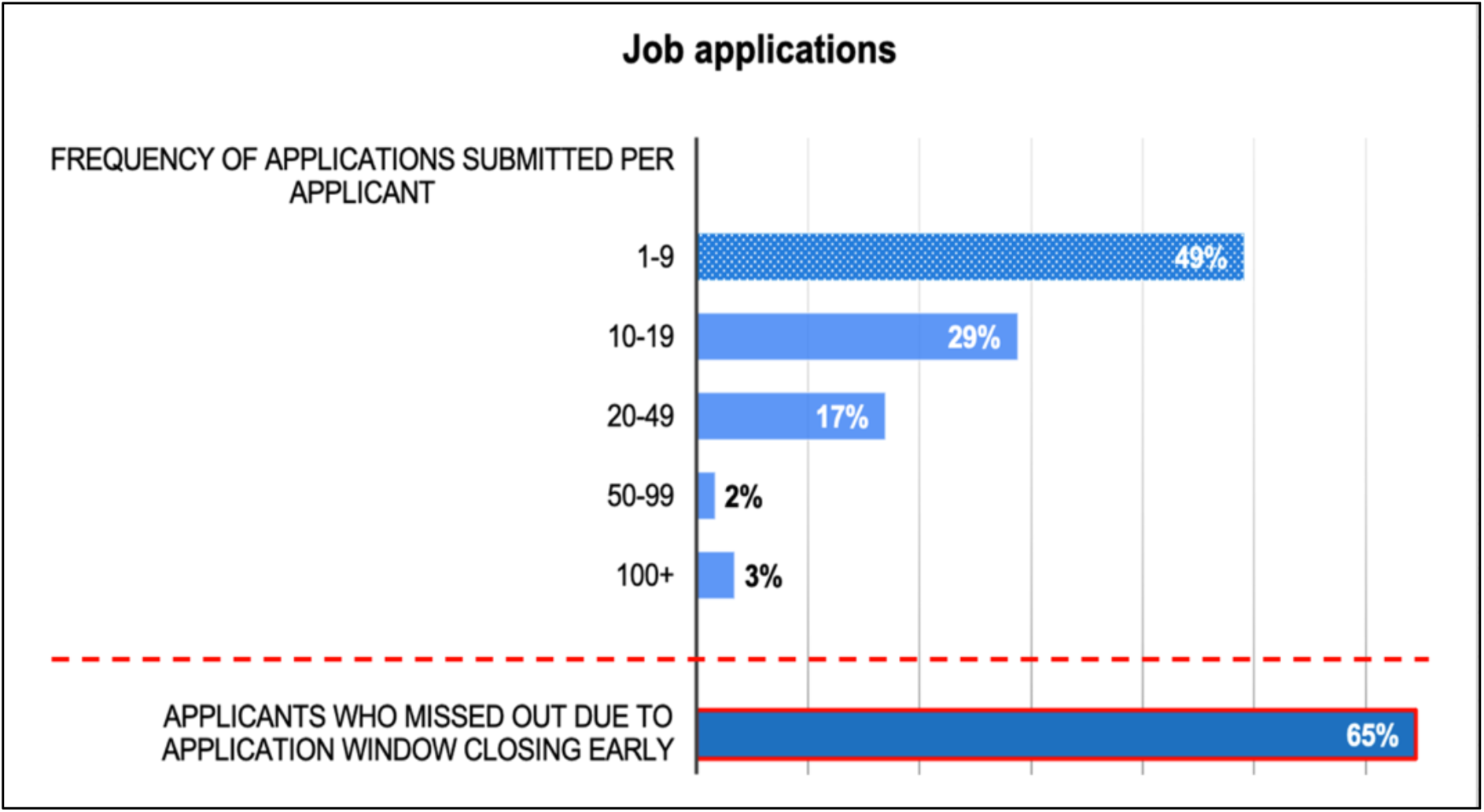
Frequency of application for local trust-based roles, 2025.

**Figure A3:**
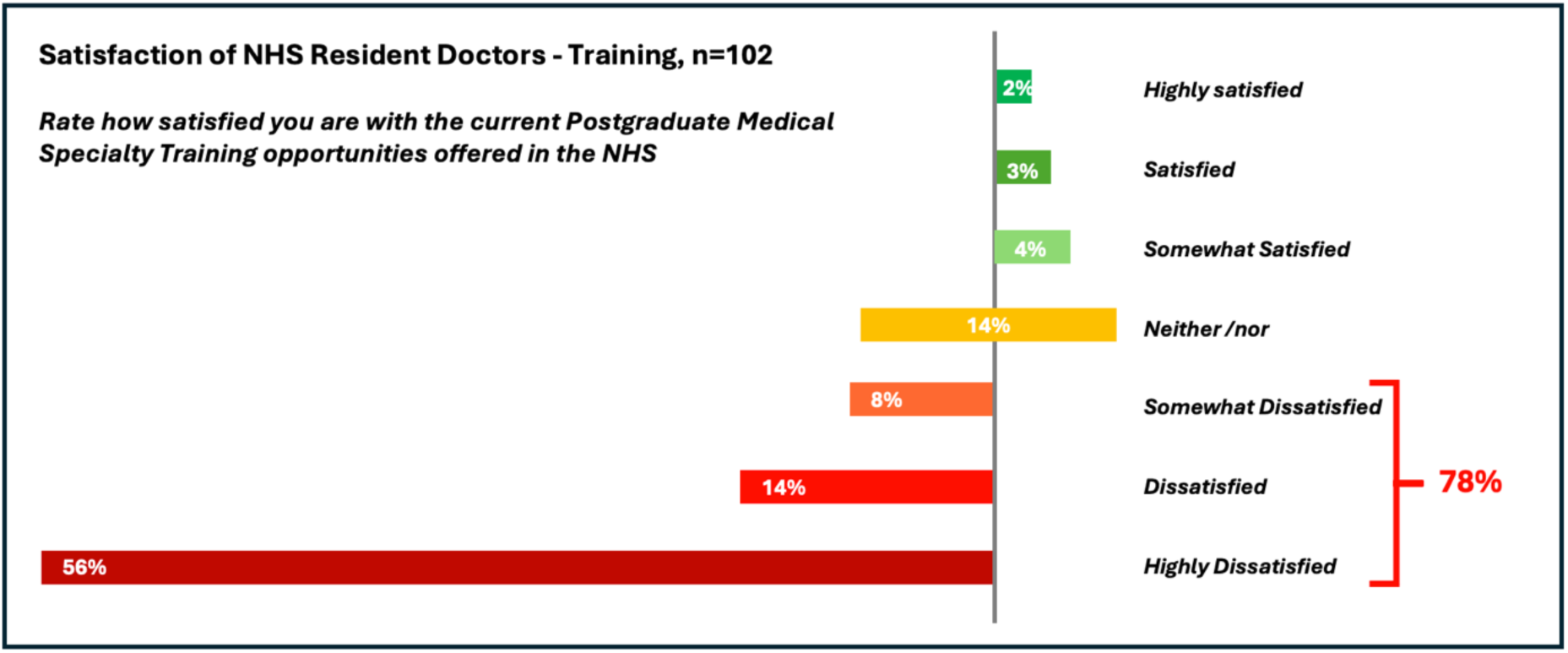
Satisfaction of NHS Resident Doctors – Specialty Training, 2025.

**Figure A4:**
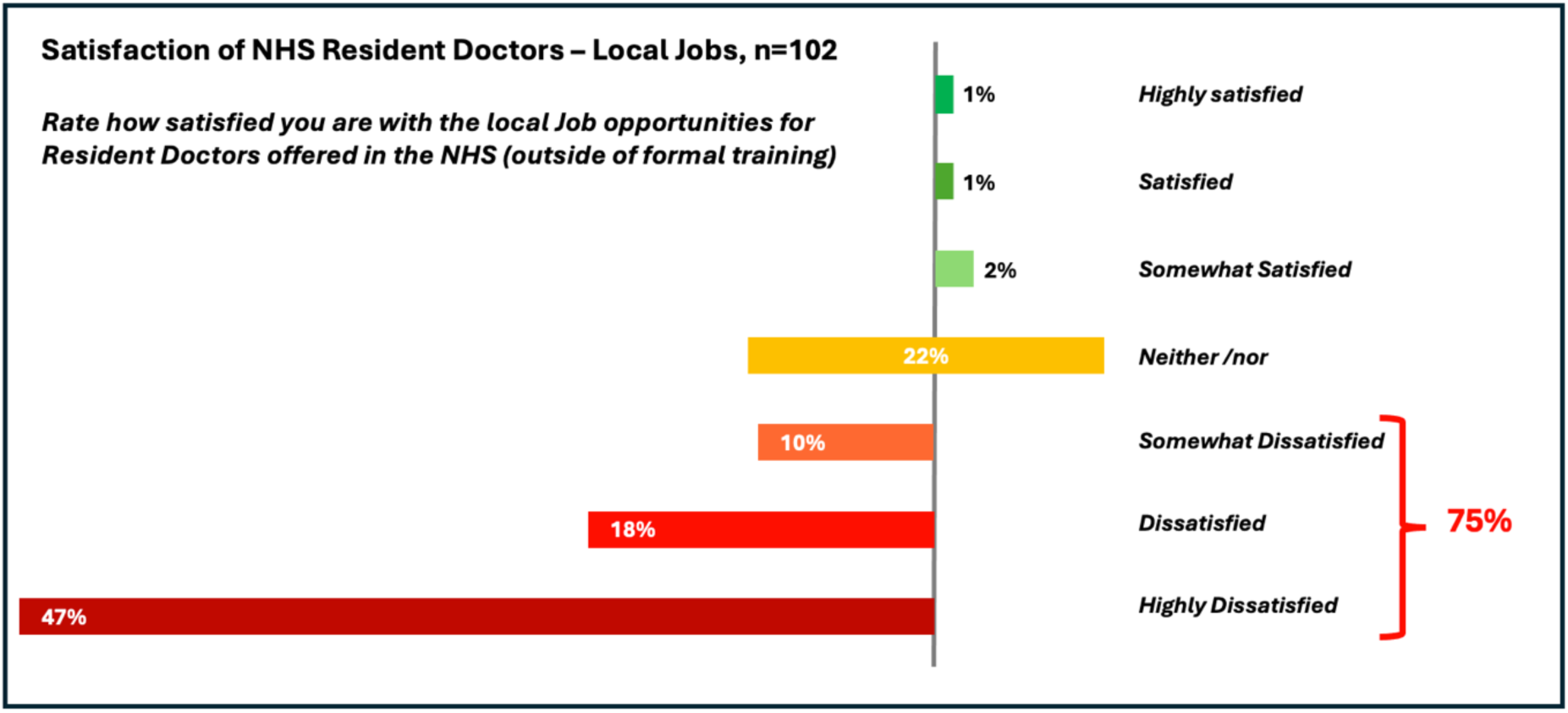
Satisfaction of NHS Resident Doctors – Local Jobs, 2025.

**Table A1:** Respondent characteristics – Level of experience.

| Year | Graduated Medical School |  | Completed Foundation Training |  |
| --- | --- | --- | --- | --- |
|  | n | % | n | % |
| ≤2017 | 6 | 6% | 6 | 6% |
| 2018 | 2 | 2% | 1 | 1% |
| 2019 | 5 | 5% | 1 | 1% |
| 2020 | 5 | 5% | 0 | 0% |
| 2021 | 11 | 11% | 5 | 5% |
| 2022 | 54 | 53% | 4 | 4% |
| 2023 | 16 | 16% | 12 | 12% |
| 2024 | 3 | 3% | 51 | 50% |
| 2025 | 0 | 0% | 18 | 18% |
| 2026 | 0 | 0% | 4 | 4% |

**Notes:** \* N= 102

**Table A2:** Thematic Insights – Representative Quotes.

| Key Theme | What doctors report | Representative Quotes |
| --- | --- | --- |
| Emotional strain and mental health | Feeling broken, emotionally exhausted and on the edge of giving up medicine; suffering deteriorating mental health. | *"I feel mentally and emotionally exhausted from trying to progress in the NHS."*<br>*"Frustrated and powerless, undervalued and left out cold, soul and confidence destroying. Had to stop applying as too detrimental for my mental health."*<br>*"Hopeless, anxious, looking into other careers."*<br>*"Scared and worried about future, helpless, trapped."*<br>*"Repeated rejections have made me feel broken, anxious, and on the edge of giving up medicine."*<br>*"It's an absolute scandal. My mental health has suffered massively, my confidence as a doctor is shattered to pieces, I won't be able to continue in medicine."* |
| Perception of being undervalued and not worthy of investment in future | Feeling undervalued and replaceable, seeing little investment in their future, and viewing medicine as a questionable career choice. | *"Undervalued. doctors are treated as numbers, not people with lives."*<br>*"I feel invisible, undervalued, and replaceable."*<br>*"Demoralised, unwanted, frustrated. Working so hard in a broken system and having to face the fact, the same system can't facilitate your ongoing training."*<br>*"It is difficult not to feel disheartened and like I have made a mistake in choosing this career."*<br>*"To be exiting F2 without a formal job despite so many applications is so discouraging and just makes me feel so ready to leave medicine now."*<br>*"Tricked, lied to, used and discarded."* |
| Job insecurity and extreme competition | Impossible to secure training posts, unrealistic competition ratios and lack of career progression. | *"It feels nearly impossible to get a training post; the process is overly competitive and demoralising."*<br>*"It's extremely disheartening to work so hard and not feel appreciated and be anxious about getting to the next stage of training."*<br>*"Undervalued, exploited, hopeless. Increased competition ratios every year with no attempt to fix it."*<br>*"It makes me feel that there is little hope for entering speciality training and very frustrated that I have invested so much time, energy and money into a career that is supposed to offer job security & career progression but has done quite the opposite."*<br>*"Uncertain, uneasy without any job security. So-called lifeline learning without life long work!"*<br>*"Frustrated, upset and feel stuck at my current level as I'm unable to progress in training."* |
| Broken by process and 'survivors' guilt | Even if successful, doctors feel demotivated and broken by the toll it took, the need to settle for less desirable opportunities and the guilt in seeing other colleagues' struggles. | *"Demoralising to see friends struggling for work, guilty that I have a training job straight out of FY2 and others are struggling for multiple years."*<br>*"I am in training but I despair for those locked out and facing unemployment."*<br>*"I have taken a CT1 surgical job that will require more than an hour commute each way. I am doing two specialities that I have no interest in pursuing further and will leave me struggling to find an ST3 post I want to train in."*<br>*"It feels bleak that despite having two Cambridge degrees and 3 years experience as a doctor working more than full time hours I am grateful to be in a job that I would have no interest in applying for outside of a national training programme."*<br>*"It was so difficult to get into ST1, and I felt so disrespected throughout the whole process I ended up turning down my offer and decided to stay in New Zealand where you feel wanted and valued and supported in your career."* |
| Systemic issues and mismanagement | A lack of workforce planning, a mismatch of roles and jobs meeting the demand from doctors, prioritisation of service provision over education. | *"Complex, multi-faceted but compounded by short-sighted workforce planning, coalescing in a minefield for all."*<br>*"Hopeless. There are rota gaps in every speciality and hospital and there are competent residents willing to do those jobs, yet the recruitment system means that those gaps go unfilled."*<br>*"Too much service provision in training posts and no training."*<br>*"Demoralised, it's sad to see people unemployed despite having understaffed working conditions."*<br>*"Rotational training is awful. Training is more about service provision. Rotas are heavy with out of hours work. Departments not taking pride in trainees."*<br>*"Training is at times fragmented and often training and teaching opportunities are missed due to poor staffing and pressures on the NHS."* |
| Overwhelmed Recruitment systems | Overburdened, failing recruitment systems, not fit for purpose and seen as unfair, untransparent and not selecting for the best candidates. | *"I have been unable to even hear back from fellow jobs when applying, yet my CV / level is perfect for some of these jobs."*<br>*"Bots are causing a lot of issues. Jobs are advertised, hundreds of applications are received, but very few if any are from people suited to the post advertised."*<br>*"Definition of madness. Does not choose the best/most suitable doctors, you have to pay to play and rewards pen pushers."*<br>*"Many changes especially in IMT recruitment over the last 6 years have meant that many resident doctors are now struggling to get into training and even risk of being unemployed, which is extremely disheartening and disappointing."* |
| Missed opportunity and inequity | Displacement of local graduates, recruitment not giving sufficient weight to relevant clinical experience, expensive upskilling requirements. | *"Bottlenecks, underfunding and political short-sightedness are threatening the future of healthcare."*<br>*"International recruitment has unintentionally displaced competent local graduates."*<br>*"Frustrated at the NHS' desperation for doctors and yet the lack of training opportunities for UK graduates. This makes no sense to me at all."*<br>*"After all this work and perceived rota gaps, nothing is being done to help the right people get in the right jobs."*<br>*"It is very costly to get to the next step through all the courses etc required to demonstrate 'commitment to specialty'."*<br>*"Too much extra-curricular non-clinical CV work required to achieve a core training place!"* |
| Exodus from the NHS and medicine | Intent to leave the NHS or the UK, migrating to countries with better conditions. Conflicted about choices and feeling forced into them. | *"Many of us are looking abroad or outside of medicine for job security, appreciation, respect, and to preserve our overall mental wellbeing."*<br>*"Desperate. I feel as if I have no choice but to leave the UK, despite wanting to stay."*<br>*"Like leaving medicine. Grad med was a total waste of time resulting in financial insecurity and stress without any job satisfaction."*<br>*"I am considering non-medical career paths but am finding it hard to fully commit to this, having trained so long to become a doctor. I also feel guilty for leaving. On the one hand, I want to help patients and fiercely believe in the NHS' core principles, but on the other, I need to protect myself as I cannot work in the NHS without it breaking me."* |

**Table A3:** Survey Tool.

| Question | Available response |
| --- | --- |
| 1. What best captures your *current* role*?<br>**\*NB:** At time when survey was conducted, 10 June and 20 July 2025 | ☐ FY1<br>☐ FY2<br>☐ In Postgraduate Specialty Training role, e.g. IMT, CT Surgery, GP Trainee, Other Specialty Trainee<br>☐ In Higher Specialty Training role (ST3 and above)<br>☐ Locally Employed Doctor, e.g. JCF, on fixed term contract (FY3 and above)<br>☐ Locally Employed Doctor, e.g. SCF, on fixed term contract (ST3 and above)<br>☐ Locum Doctor - SHO/FY3 and above (full time Locum Doctor only, not in addition to other fixed term contract or training posts)<br>☐ Locum Doctor - SpR/ST3 and above (full time Locum Doctor only, not in addition to other fixed term contract or training posts)<br>☐ Working within the NHS, but in a non-clinical or mixed role, such as Research or Teaching Fellow<br>☐ Working in private sector in the UK (as a doctor)<br>☐ Working in private sector in the UK (in a non-clinical role)<br>☐ Working abroad as a doctor<br>☐ Working abroad in a non-clinical role<br>☐ Other: _____ |
| 2. Please select what best describes your role *from August 2025* (either confirmed or most likely) | ☐ FY1<br>☐ FY2<br>☐ In Postgraduate Specialty Training role, e.g. IMT, CT Surgery, GP Trainee, Other Specialty Trainee<br>☐ In Higher Specialty Training role (ST3 and above)<br>☐ Locally Employed Doctor, e.g. JCF, on fixed term contract (FY3 and above)<br>☐ Locally Employed Doctor, e.g. SCF, on fixed term contract (ST3 and above)<br>☐ Locum Doctor - SHO/FY3 and above (full time Locum Doctor only, not in addition to other fixed term contract or training posts)<br>☐ Locum Doctor - SpR/ST3 and above (full time Locum Doctor only, not in addition to other fixed term contract or training posts)<br>☐ Working within the NHS, but in a non-clinical or mixed role, such as Research or Teaching Fellow<br>☐ Working in private sector in the UK (as a doctor)<br>☐ Working in private sector in the UK (in a non-clinical role)<br>☐ Working abroad as a doctor<br>☐ Working abroad in a non-clinical role<br>☐ Other: _____ |
| 3. Have you ever applied to any of the NHS specialty training programs outside of the Foundation Program? | If YES, please indicate how many training programs in total you have applied for over the years/recruitment cycles.<br>• Answer: _____<br>If NO, go straight to Question 10 |
| 4. Have you applied to any specialty training programs for **entry in 2025**? | If YES, tick all that apply<br>☐ IMT1<br>☐ IMT3<br>☐ Surgery Core CT1<br>☐ Psychiatry Core CT1<br>☐ ACCS/Anaesthetics CT1<br>☐ GP Training ST1<br>☐ Paediatrics ST1<br>☐ Obs and Gynae ST1<br>☐ Other Run Through Programmes (ST1)<br>☐ Higher Specialty Training (ST3 and above)<br>☐ Other: _____<br>If NO, go to Question 7 |
| 5. If YES please indicate how many applications resulted in shortlisting/ interviews (for entry in **2025**) | • Answer: _____ |
| 6. If YES and you have been shortlisted / interviewed, please indicate how many resulted in a training place offer (for entry in **2025**). | • Answer: _____ |
| 7. Did you apply to any specialty training programs for **entry in 2024**? | If YES, tick all that apply<br>☐ IMT1<br>☐ IMT3<br>☐ Surgery Core CT1<br>☐ Psychiatry Core CT1<br>☐ ACCS/Anaesthetics CT1<br>☐ GP Training ST1<br>☐ Paediatrics ST1<br>☐ Obs and Gynae ST1<br>☐ Other Run Through Programmes (ST1)<br>☐ Higher Specialty Training (ST3 and above)<br>☐ Other: _____<br>If NO, go to Question 10 |
| 8. If YES please indicate how many applications resulted in shortlisting/ interviews (for entry in **2024**) | • Answer: _____ |
| 9. If YES and you have been shortlisted / interviewed, please indicate how many resulted in a training place offer (for entry in **2024**). | • Answer: _____ |
| 10. Have you ever applied or tried to apply for a NHS fixed term contract or permanent job, e.g. LED, JCF or SCF roles or Trust Grade Jobs? | ☐ Yes<br>☐ No. Go to Question 15 |
| 11. If YES, please fill in the number of jobs you were interested in (or closest estimate), but for which you could **NOT** submit an application, as the job application window closed early / before the advertised deadline | • Answer: _____ |
| 12. If YES please fill in the total number of applications you have submitted (or closest estimate) | • Answer: _____ |
| 13. If YES please fill in how many (or closest estimate) of these submitted applications resulted in an interview/ shortlisting | • Answer: _____ |
| 14. If YES and you have been interviewed/shortlisted for a job, please indicate how many resulted in a job offer. Otherwise leave blank. | • Answer: _____ |
| 15. Rate how satisfied you are with the current Postgraduate Medical Specialty Training opportunities offered in the NHS. | 1 2 3 4 5 6 7 8 9 10<br>Not satisfied at all ☐ ☐ ☐ ☐ ☐ ☐ ☐ ☐ ☐ ☐ Highly satisfied |
| 16. Rate how satisfied you are with the current Resident Doctor Job opportunities (outside of training programs) offered in the NHS | 1 2 3 4 5 6 7 8 9 10<br>Not satisfied at all ☐ ☐ ☐ ☐ ☐ ☐ ☐ ☐ ☐ ☐ Highly satisfied |
| 17. Please describe in a few words how the current Postgraduate Medical Specialty Training and wider resident doctor job situation in the NHS makes you feel | • Answer: _____ |
| Please provide some further info about your Medical Training (if you want to) and select as applicable. | ☐ I am a UK Medical School Graduate (UK national)<br>☐ I am a UK Medical School Graduate (non-UK national)<br>☐ I am a International Medical Graduate (UK national)<br>☐ I am a International Medical Graduate (non-UK national)<br>☐ Other: _____ |
| What year did you graduate from Medical School? | Answer: _____ |
| When have you or will you complete your Foundation Training / FY2 ? | Answer: _____ |

**Table A4:** The STROBE reporting checklist.

|  | Item Description | Location (or reason for not reporting) |
| --- | --- | --- |
| **Title and abstract** |  |  |
| 1a. Indicate the study's design | Indicate the study's design with a commonly used term in the title or the abstract. | Page 1, Title |
| 1b. Abstract | Provide in the abstract an informative and balanced summary of what was done and what was found. | Page 1, Abstract |
| **Introduction** |  |  |
| 2. Background / rationale | Explain the scientific background and rationale for the investigation being reported. | Page 2, Introduction |
| 3. Objectives | State specific objectives, including any prespecified hypotheses. | Page 2, included in Introduction |
| **Methods** |  |  |
| 4. Study design | Present key elements of study design early in the paper. | Page 3, Methods |
| 5. Setting | Describe the setting, locations, and relevant dates, including periods of recruitment, exposure, follow-up, and data collection. | Page 3, Methods |
| 6a. Eligibility criteria | **Cohort study:** Give the eligibility criteria, and the sources and methods of selection of participants. Describe methods of follow-up.<br>**Case-control study:** Give the eligibility criteria, and the sources and methods of case ascertainment and control selection. Give the rationale for the choice of cases and controls.<br>**Cross-sectional study:** Give the eligibility criteria, and the sources and methods of selection of participants. | Page 3, Methods |
| <u>6b. Matching criteria</u> | **Cohort study:** For matched studies, give matching criteria and number of exposed and unexposed. **Case-control study:** For matched studies, give matching criteria and the number of controls per case. | n/a |
| <u>7. Variables</u> | Clearly define all outcomes, exposures, predictors, potential confounders, and effect modifiers. Give diagnostic criteria, if applicable. | n/a |
| <u>8. Data sources / measurement</u> | For each variable of interest give sources of data and details of methods of assessment (measurement). Describe comparability of assessment methods if there is more than one group. | n/a |
| <u>9. Bias</u> | Describe any efforts to address potential sources of bias. | Page 3, Methods |
| <u>10. Study size</u> | Explain how the study size was arrived at. | n/a |
| <u>11. Quantitative variables</u> | Explain how quantitative variables were handled in the analyses. If applicable, describe which groupings were chosen, and why. | Page 3, Methods |
| <u>12a. Statistical methods</u> | Describe all statistical methods, including those used to control for confounding. | n/a |
| <u>12b. Statistical methods – subgroups and interactions</u> | Describe any methods used to examine subgroups and interactions. | n/a |
| <u>12c. Statistical methods – missing data</u> | Explain how missing data were addressed. | Page 3, Methods |
| <u>12di. Statistical methods – loss to follow-up</u> | **Cohort study:** If applicable, describe how loss to follow-up was addressed. | n/a |
| <u>12dii. Statistical methods – matching cases and controls</u> | **Case-control study:** If applicable, explain how matching of cases and controls was addressed. | n/a |
| <u>12diii. Statistical methods – sampling strategy</u> | **Cross-sectional study:** If applicable, describe analytical methods taking account of sampling strategy. | Page 3, Methods |
| <u>12e. Statistical methods – sensitivity analyses</u> | Describe any sensitivity analyses. | n/a |
| **Results** |  |  |
| 13a. Participant numbers | Report the numbers of individuals at each stage of the study—e.g., numbers potentially eligible, examined for eligibility, confirmed eligible, included in the study, completing follow-up, and analysed; Consider use of a flow diagram. | Page 4, Results |
| 13b. Participants – non-participation | Give reasons for non-participation at each stage. | n/a |
| 13c. Participants – flow diagram | Consider use of a flow diagram. | n/a |
| 14a. Descriptive data – participant characteristics | Give characteristics of study participants (e.g., demographic, clinical, social) and information on exposures and potential confounders. Present the information in a table. | Page 4, Results |
| 14b. Descriptive data – missing data | Indicate the number of participants with missing data for each variable of interest. | Page 4, Results |
| 14c. Descriptive data – follow-up time | **Cohort study:** Summarise follow-up time—e.g., average and total amount. | n/a |
| 15. Outcome data | **Cohort study:** Report numbers of outcome events or summary measures over time. **Case-control study:** Report numbers in each exposure category, or summary measures of exposure. **Cross-sectional study:** Report numbers of outcome events or summary measures. | Page 4, Results |
| 16a. Main results | Give unadjusted estimates and, if applicable, confounder-adjusted estimates and their precision (e.g., 95% confidence intervals). Make clear which confounders were adjusted for and why they were included. | n/a |
| 16b. Main results – category boundaries | Report category boundaries when continuous variables were categorised. | n/a |
| 16c. Main results – risk | If relevant, consider translating estimates of relative risk into absolute risk for a meaningful time period. | n/a |
| 17. Other analyses | Report other analyses done—e.g., analyses of subgroups and interactions, and sensitivity analyses. | Page 6, Qualitative Sentiment Analysis |
| **Discussion** |  |  |
| 18. Key results | Summarise key results with reference to study objectives. | Page 7, Discussion |
| 19. Limitations | Discuss limitations of the study, taking into account sources of potential bias or | Page 2, Article Summary |
|  | imprecision. Discuss both direction and magnitude of any potential bias. |  |
| <u>20. Interpretation</u> | Give a cautious overall interpretation considering objectives, limitations, multiplicity of analyses, results from similar studies, and other relevant evidence. | Page 7, Discussion |
| <u>21. Generalisability</u> | Discuss the generalisability (external validity) of the study results. | Page 7, Discussion |
| **Other information** |  |  |
| <u>22. Funding</u> | Give the source of funding and the role of the funders for the present study and, if applicable, for the original study on which the present article is based. | Page 9 |

